# Family dynamics and environmental factors influencing progression from alcohol experimentation to initiation in youth: Evidence from the Adolescent Brain Cognitive Development (ABCD) Study

**DOI:** 10.1101/2025.06.23.25330149

**Authors:** Maya I. Harwood, Daniel F. Otero-Leon, Erin J. Stringfellow, Arielle R. Deutsch, Mohammad S. Jalali, Huiru Dong

## Abstract

**Purpose:** While a growing body of research categorizes early drinking behaviors into experimentation and initiation, factors driving the transition remain underexplored. This study examined individual and environmental influences on the time from alcohol experimentation to initiation among preadolescent youth.

**Methods:** Data came from the Adolescent Brain Cognitive Development (ABCD) Study (2016-2021). Experimentation was defined as first sip and initiation as first full drink during follow-up. Extended Cox models assessed the impact of sociodemographic factors, alcohol expectancies, family and peer dynamics, and neighborhood-level factors on the likelihood of alcohol initiation following experimentation.

**Results:** Of 1,213 youths, 87 (7.2%) progressed from sipping to drinking within 45 months. Older age at onset of sipping was associated with the highest likelihood of alcohol initiation (adjusted hazard ratio [HR]=6.40, 95% confidence interval [CI]: 3.72-11.02, p<0.001), followed by peer alcohol use (adjusted HR=4.11, 95% CI: 2.55-6.65, p<0.001), household rules allowing alcohol consumption (adjusted HR=2.14, 95% CI: 1.17-3.91, p=0.014), family conflict (adjusted HR=1.14, 95% CI: 1.04-1.26, p=0.007), and positive alcohol expectancies (adjusted HR=1.10, 95% CI: 1.01-1.21, p=0.032). Conversely, older chronological age was protective (adjusted HR=0.30, 95% CI: 0.18-0.51, p<0.001). No significant differences in the likelihood of alcohol initiation were observed based on sex, race, ethnicity, household income, parental drinking problems, alcohol availability, and neighborhood safety.

**Discussion:** Age, social and familial influences, and positive alcohol expectancies play central roles in early drinking progression. Interventions that integrate individual beliefs, family practices, and peer dynamics should be considered for delaying initiation.

## INTRODUCTION

Preadolescence and adolescence are critical stages for the onset of alcohol use. In 2024, 18.5% of eighth graders in the U.S. reported having consumed alcohol at least once in their lifetime, a figure that rose to 32% among tenth graders [1]. Early onset of alcohol consumption is linked to the later development of problem drinking and other negative health outcomes, including increased risk of suicidality and changes in personality traits [2, 3, 4, 5]. Given the developmental sensitivity of preadolescence, identifying risk and protective factors for alcohol use among youth is crucial to guiding prevention efforts.

Preadolescent alcohol experimentation (i.e., sipping) is increasingly recognized as a qualitatively distinct stage compared to alcohol initiation (i.e., consuming a full drink) in the developmental trajectory of youth alcohol use. Alcohol sipping, often defined as tasting or having a few sips of alcohol [6, 7] commonly precedes initiation, which signals a shift toward more intentional use and greater vulnerability to future negative health outcomes [8]. While previous research has made significant strides in identifying factors influencing youth alcohol use, the transition from sipping to consuming a full drink remains underexplored.

Studies show that sipping is far more prevalent than initiation with a full drink during preadolescence and is strongly shaped by parental modeling and approval [9]. Building on this, other research shows that early sippers, compared to youth who have consumed full drinks, tend to come from more restrictive environments, marked by stricter parental monitoring and lower exposure to substance-using peers [10]. These findings suggest that sippers may face fewer environmental opportunities to initiate drinking, even as they begin to experiment. Paradoxically, early sippers also exhibit characteristics that are typically associated with greater risk for externalizing behavior and substance use, including heightened sensation seeking, disinhibition, and weaker negative alcohol expectancies [11, 12]. This contradiction – behavioral vulnerability within socially restrictive contexts – distinguishes them from early initiators of full drinks, who are more consistently linked to permissive parental attitudes, easier access to alcohol, and affiliation with substance-using peers [13, 14].

Together, these findings highlight critical gaps in understanding what drives the transition from sipping to initiation. Our study builds on this foundation by examining how individual-level traits, parental attitudes, and peer behavior jointly shape the progression from alcohol sipping to initiation. Understanding sipping as a unique point along the alcohol use continuum, rather than equating it with abstinence or drinking, may offer new insights into early prevention opportunities before initiation occurs.

Additionally, few longitudinal cohort studies on alcohol use have focused specifically on preadolescent youth, despite this being a developmentally sensitive period when early experimentation often begins [15]. Preadolescence differs from later adolescence in critical ways, including more limited autonomy and stronger parental influence [16]. These distinctions suggest that the factors influencing early alcohol use trajectories may differ in both nature and timing compared to older youth. However, little is known about how youths’ relationships and environment interact to influence the progression from initial experimentation to initiation during this stage. This gap in the literature underscores the need for research that captures the early and nuanced dynamics of alcohol use development in childhood.

This study leverages the Adolescent Brain Cognitive Development (ABCD) Study, a longitudinal cohort of over 11,000 youth followed from ages 9-10 into early adulthood [17], to examine the progression from alcohol experimentation (sipping) to initiation (first full drink). While prior ABCD-based studies have focused separately on the onset of either sipping or a full drink, none have addressed the transition between these distinct stages [18, 19]. By drawing on detailed measures of time-updated peer behavior, household dynamics, and alcohol expectancies, we aim to identify individual, interpersonal, and environmental factors that shape the timing of this progression toward more intentional and potentially risky drinking behaviors.

## METHODS

### Study participants

The ABCD Study tracks the biological and behavioral development of 11,868 participants recruited at ages 9-10 across 17 US states [17]. We used data from the ABCD 5.1 release, which was collected from 2016-2021 with follow-ups occurring every six months, with ages ranging from 8 to 11 at baseline and 12 to 14 at the end of the study period [17]. For each participant, alcohol consumption was assessed during annual in-person interviews and mid-year phone interviews. Alcohol experimentation was defined as the first instance of alcohol sipping, while alcohol initiation was defined as consuming at least one standard drink containing 0.6 fluid ounces or 14 grams of pure alcohol [20]. Since the study aims to identify factors associated with alcohol initiation following experimentation, the inclusion criteria consisted of no alcohol use at baseline and engaging in experimentation (alcohol sipping) at least once during the study. Additionally, to establish a temporal relationship from alcohol experimentation to initiation, only participants with complete and chronological interview dates were included. Furthermore, participants with no follow-up after reporting their first sip were excluded. Lastly, among participants who sipped alcohol and had a full drink, only those who reported sipping prior to consuming a full drink were retained. Figure 1 depicts the process used to determine our final cohort of 1,213 participants. See Table A1 for participant characteristics of those excluded in Boxes B, C, and D.

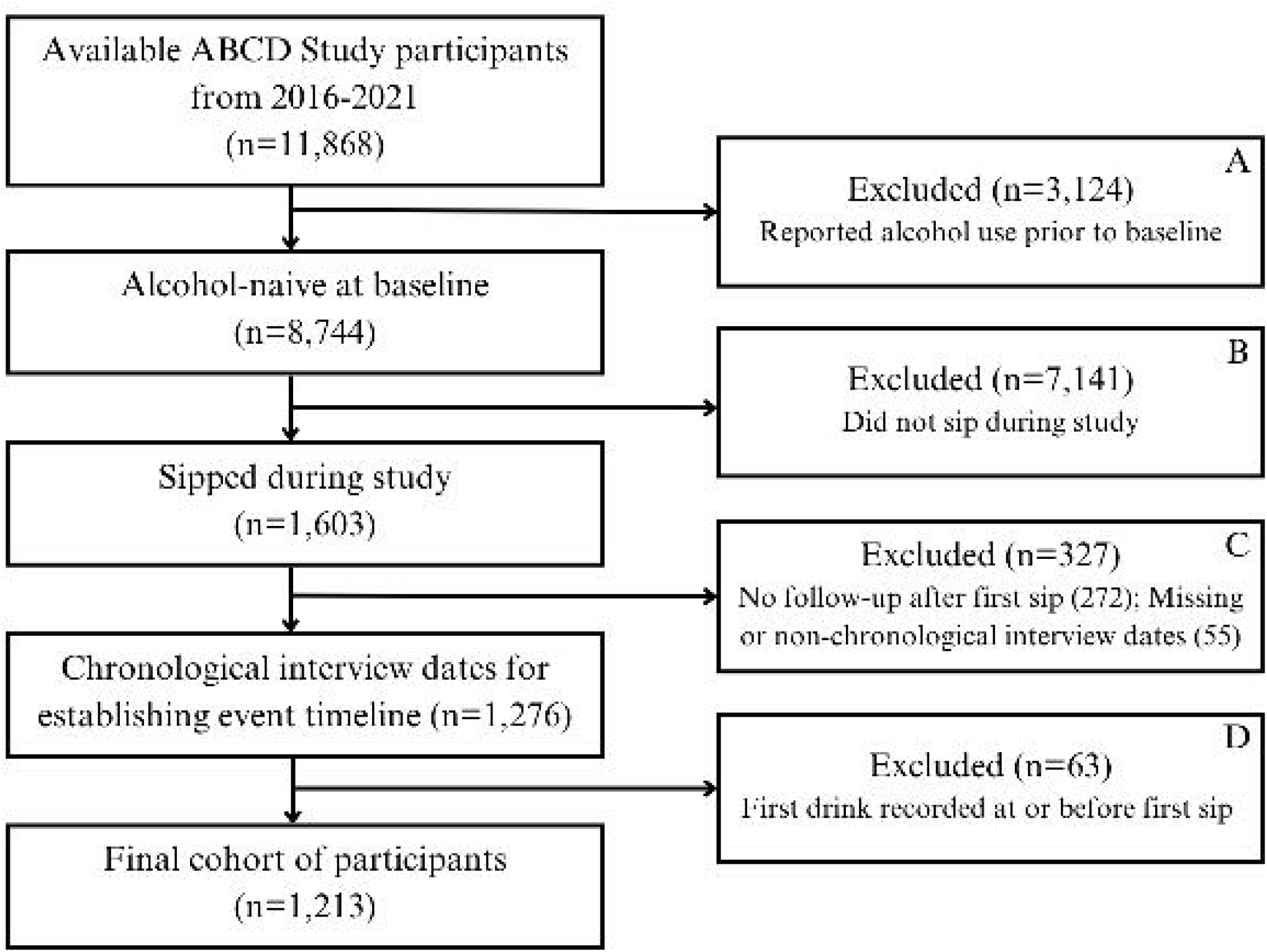

### Measures

The outcome is youth self-reported alcohol initiation, assessed by the question: “Have you had a full drink containing alcohol at any time in the past 6 months?” [20]. We included thirteen explanatory factors spanning individual, interpersonal, and environmental aspects. Covariates were chosen based on established or plausible association with alcohol use [21, 22, 23], with a focus on measures of family, household, and peer dynamics, to holistically assess the risk of alcohol initiation following experimentation.

Specifically, we considered six sociodemographic characteristics, including time-updated age, age at onset of sipping, sex (male; female), race (White; Black; multiracial or other), ethnicity (Hispanic; non-Hispanic), and baseline household income (<$50,000; $50,000-$100,000; >$100,000) reported by a parent or guardian. Peer alcohol use, defined as whether a youth’s friends consume alcohol and collected as part of a nine-item Peer Group Deviance instrument modified from the PhenX, was reported by youth on a yearly basis [24]. Family dynamics were assessed using several measures reported by a parent or guardian. This included parental drinking problems (yes or no for each parent), which was collected at baseline. Household rules regarding youth’s alcohol consumption (alcohol consumption allowed; alcohol consumption not allowed; no rules on alcohol consumption in the home) and alcohol availability in the home (easy; difficult) were collected once a year. Additionally, family conflict was measured using the Family Environment Scale-Conflict Subscale (FES-CS) modified from the PhenX, a nine-item instrument capturing the amount of openly expressed anger and conflict in a family [25]. Scores range from 1 to 9, with higher scores indicating more frequent expressions of anger and interactions that stimulate anger.

Moreover, we included positive and negative alcohol expectancies reported by youth once a year. Alcohol expectancies were assessed using the Alcohol Expectancy Questionnaire-Adolescent, Brief, a seven-item questionnaire capturing both positive (e.g., alcohol helps a person relax) and negative (e.g., alcohol leads to being mean to others) expectations regarding alcohol’s effects [26]. Participants indicated their level of agreement with statements regarding alcohol’s effects using a 7-point Likert scale (1 = “strongly disagree” to 5 = “strongly agree”). A positive alcohol expectancy score was derived by summing responses to items 1, 2, 4, and 6, while a negative alcohol expectancy score was obtained by summing responses to items 3, 5, and 7. Lastly, neighborhood safety, which was collected from a parent or guardian once a year, was measured using a 3-item Neighborhood Safety instrument modified from the PhenX that produces scores ranging from 1 to 5 [27].

### Analysis

First, we examined the characteristics of the ABCD Study participants and stratified them by alcohol initiation, the outcome of interest. They were further compared using chi-squared tests for categorical variables (except for race, peer alcohol use, and household alcohol consumption rules, which were compared using Fisher’s exact tests) and t-tests for continuous variables. To visualize the cumulative probability of initiating alcohol use after experimentation, we constructed a cumulative incidence curve. Then, bivariable and multivariable extended Cox models were used to assess the influence of sociodemographic characteristics, family dynamics, peer alcohol use, alcohol expectancies, and neighborhood-level factors on the transition from alcohol experimentation to initiation. Unlike a standard Cox model, this approach relaxed the proportional hazards assumption and enabled the inclusion of time-dependent covariates such as age, alcohol expectancies, and peer alcohol use [28]. To assess whether the effects of these factors varied as participants aged, we conducted a secondary analysis testing their interactions with age. Participants who had not initiated drinking by the end of the study period were censored at their last recorded observation.

Statistical analyses were conducted in R (version 4.1.2) using the *coxph* function in the *survival* package to fit the extended Cox model. All p-values were two-sided with the significance level as 0.05. All analytical codes can be found at 10.5281/zenodo.16929131. The present study is a secondary analysis of de-identified ABCD Study data and was deemed exempt from informed consent by the Massachusetts General Hospital institutional review board.

## RESULTS

Table 1 summarizes key baseline characteristics of alcohol-naive participants who reported sipping alcohol during the study. Among 1,213 youths in the final sample, 87 (7.2%) participants had their first drink after taking their first sip during the study period, as shown in Figure 2. Participants who initiated alcohol use were slightly older at baseline, with no significant differences in sex, race, ethnicity, or household income. A comparison of the final included participants with excluded participants is provided in the appendix.

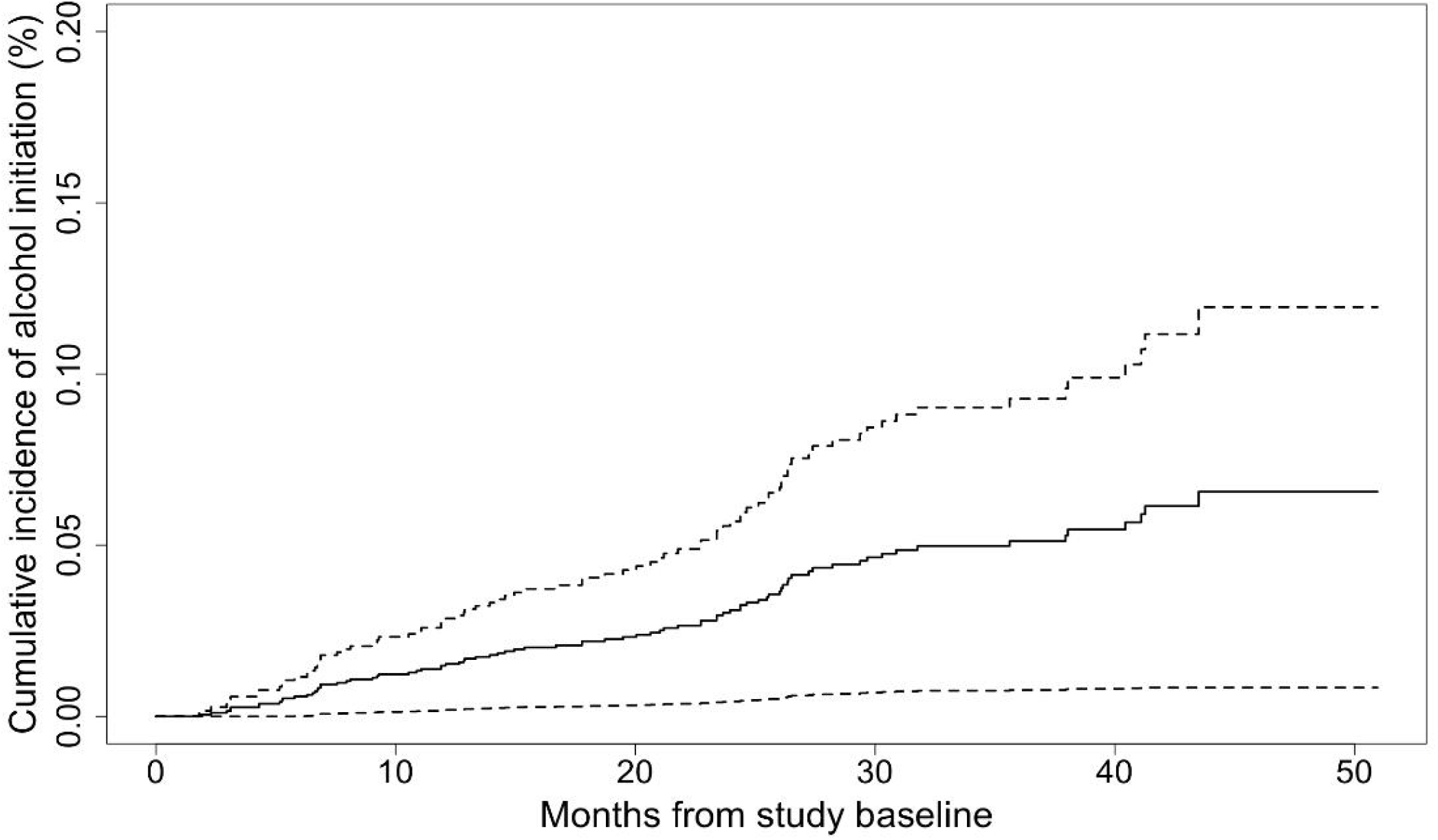

As shown in Table 2, several peer and family dynamics were significantly associated with alcohol use progression. Specifically, peer alcohol use emerged as a strong predictor of alcohol initiation. Youth who reported having friends who consumed alcohol were over four times likely to initiate drinking themselves compared to those whose peers did not drink (adjusted hazard ratio [HR] = 4.11, 95% CI: 2.55–6.65). Household rules also played a significant role. Compared to youth living in homes where alcohol consumption was not allowed, those whose parents allowed alcohol use were at a 114% higher risk of initiating alcohol (adjusted HR = 2.14, 95% CI: 1.17–3.91). Family conflict was also positively associated with the time from alcohol experimentation to initiation. For each unit increase in family conflict score, the hazard of initiation increased by 14% (adjusted HR = 1.14, 95% CI: 1.04–1.26).

Sociodemographic factors, such as sex, race, ethnicity, and household income, were not significantly associated with alcohol initiation in these study participants after adjusting for other variables. Older age at onset of sipping was associated with a higher likelihood of progressing from experimentation to initiation (adjusted HR = 6.40, 95% CI: 3.72–11.02). Conversely, each additional year of chronological age was linked to a 70% lower likelihood of progression (adjusted HR = 0.30, 95% CI: 0.18–0.51). In the secondary analysis, there was no evidence that the effects of covariates were moderated by age.

Positive alcohol expectancies were modestly associated with alcohol initiation, suggesting that a youth’s perception of potential benefits from drinking may increase their likelihood of progressing beyond experimentation (adjusted HR = 1.10, 95% CI: 1.01–1.21). In contrast, negative alcohol expectancies did not show a significant protective effect, indicating that simply recognizing the risks of alcohol use may not be enough to deter initiation.

## DISCUSSION

This study examined the transition from alcohol experimentation to initiation in a large, national sample of preadolescent youth in the U.S. Using data from the ABCD Study, we identified several key factors linked to this transition, including peer alcohol use, permissive household rules, older age at onset of sipping, family conflict, and positive alcohol expectancies. In contrast, parental drinking problems, alcohol availability, and negative alcohol expectancies were not significantly associated with initiation.

Among sociodemographic variables, age at onset of sipping and chronological age were significantly associated with initiation risk. Specifically, delaying sipping by one year was linked to more than six times the hazard of transitioning from experimentation to initiation. Older chronological age at any given point was associated with a substantially lower hazard of progression, suggesting that age-related developmental factors may provide a protective effect. Together, the results indicate that the timing of first exposure relative to a youth’s developmental stage may be more important than chronological age in determining progression risk. In contrast, sex, race, ethnicity, and household income were not significant explanatory factors in the adjusted model.

Peer alcohol use emerged as a strong predictor of progression to alcohol initiation. Youth who reported having friends who drank were four times likely to transition from sipping to consuming a full drink. While peer influence has long been recognized as a risk factor, few studies directly account for the effects of peer versus parental behavior in the same model. Our findings suggest that during preadolescence and early adolescence, peer drinking habits may have a more immediate and powerful influence on drinking progression from sipping to full drink than parental modeling. Notably, parental drinking problems did not significantly predict initiation, diverging from past research that highlights parental modeling as a key risk factor [29]. It is possible that parental drinking exerts its influence over longer-term behavioral trajectories or interacts with other parenting factors, such as rule-setting, that were separately accounted for in our model. Future studies should further explore these potential mechanisms.

Family dynamics influenced alcohol use progression in youth. Permissive household alcohol rules were significantly associated with increased initiation risk. Youth in homes where alcohol consumption was allowed were 114% more likely to initiate use compared to those in homes with no-use rules. This is consistent with prior findings that permissive parenting styles around substances contribute to riskier use [30]. Additionally, family conflict was positively associated with alcohol initiation. Youth who reported greater levels of conflict in the home had a higher likelihood of progressing to initiation. This aligns with prior studies that link high-conflict family environments to increased substance use in adolescence [31]. Conflict may reduce parental monitoring or increase emotional distress, both of which can contribute to early alcohol use.

Our findings regarding alcohol expectancies provide further insight into the psychological mechanisms underlying initiation. Positive alcohol expectancies were significantly associated with increased initiation risk, while negative alcohol expectancies were not. This finding indicates that perceived benefits often exert a stronger motivational pull than perceived risks. Moreover, Sanchez et al. [32] demonstrated that family conflict and peer victimization predicted higher positive alcohol expectancies among adolescents, reinforcing the idea that environmental stressors may contribute to both risk perception and alcohol-related decision-making. These findings suggest that interventions aiming to delay initiation should not only correct misinformation about alcohol’s effects but also address the environmental and interpersonal contexts that shape these beliefs.

Neighborhood safety and alcohol availability were not significantly associated with initiation in our analysis. The null effect of neighborhood safety on initiation is consistent with other findings from ABCD Study data that reveal no association between neighborhood crime rates and substance initiation [33]. However, our alcohol availability finding contrasts with earlier studies showing that increased alcohol availability in the home correlates with earlier use [14]. One possibility is that in a younger age group, direct access to alcohol is more tightly controlled by parents, making household rules more immediate determinants of behavior.

Despite leveraging a rich longitudinal dataset, this study has limitations. The ABCD Study sample is not fully representative of the U.S. population; participants tend to come from families with above-average income and education levels [34], potentially biasing observed risk estimates. Additionally, the six-month interval between assessments may have reduced the precision in capturing the exact timing of alcohol initiation. This possibility limited the ability to identify transitions that occurred between visits. Lastly, the current study focused on socio-demographics, family and peer dynamics, alcohol expectancies, and environmental factors. Expanding the range of variables included in the analysis could deepen our understanding of the mechanisms underlying early substance use.

In conclusion, this study highlights the critical role of age, peers, family dynamics, and positive alcohol expectancies in influencing youth alcohol initiation. By focusing specifically on the transition from experimentation to initiation and leveraging time-updated covariates in a longitudinal model, our findings offer timely insights for prevention strategies that involve family and peer networks to delay or prevent alcohol use progression in at-risk youth.

## Data Availability

All data produced are available from the ABCD Study via the NIMH Data Archive.

