## Supplemental Table 1 for "Family dynamics and environmental factors influencing progression from alcohol experimentation to initiation in youth: Evidence from the Adolescent Brain Cognitive Development (ABCD) Study"

Supplementary Table S1: Baseline characteristics of three excluded groups (displayed in Boxes B (N=7,141), C (N=327), and D (N=63) in Figure 1) compared to baseline characteristics of our final sample of 1,213 ABCD Study participants from 2016-2021 (displayed in Table 1).

|  | Excluded in Box B of Figure 1 (N=7,141) | | Excluded in Box C of Figure 1 (N=327) | | Excluded in Box D of Figure 1 (N=63) | |
| --- | --- | --- | --- | --- | --- | --- |
|  |  | *p* |  | *p* |  | *p* |
| **Age** |  | <0.001 |  | 0.746 |  | 0.071 |
| Mean (SD) | 9.45 (0.54) |  | 9.56 (0.55) |  | 9.63 (0.55) |  |
| **Sex** |  | 0.057† |  | <0.001† |  | 0.001† |
| Male | 3,627 (50.8%) |  | 139 (42.5%) |  | 21 (33.3%) |  |
| Female | 3,513 (49.2%) |  | 188 (57.5%) |  | 42 (66.7%) |  |
| Intersex Male | 1 (0.0%) |  | 0 (0.0%) |  | 0 (0.0%) |  |
| **Race** |  | <0.001 |  | 0.373 |  | 0.762 |
| White | 4,281 (59.9%) |  | 223 (68.2%) |  | 47 (74.6%) |  |
| Black | 1,325 (18.6%) |  | 38 (11.6%) |  | 6 (9.5%) |  |
| Multiracial or Other | 1,415 (19.8%) |  | 62 (19.0%) |  | 10 (15.9%) |  |
| **Ethnicity** |  | 0.078 |  | 0.326 |  | 0.353 |
| Non-Hispanic | 5,571 (78.0%) |  | 256 (78.3%) |  | 47 (74.6%) |  |
| Hispanic | 1,471 (20.6%) |  | 69 (21.1%) |  | 15 (23.8%) |  |
| **Income** |  | <0.001 |  | 0.367 |  | 0.004 |
| <$50,000 | 2,126 (29.8%) |  | 75 (22.9%) |  | 25 (39.7%) |  |
| $50,000-100,000 | 1,911 (26.8%) |  | 91 (27.8%) |  | 15 (23.8%) |  |
| >$100,000 | 2,469 (34.6%) |  | 139 (42.5%) |  | 22 (34.9%) |  |
| **Maternal Drinking Problem** |  | 0.007 |  | <0.001 |  | 0.196† |
| No | 6,603 (92.5%) |  | 294 (89.9%) |  | 57 (90.5%) |  |
| Yes | 318 (4.5%) |  | 22 (6.7%) |  | 4 (6.3%) |  |
| **Paternal Drinking Problem** |  | 0.728 |  | 0.730 |  | 0.341 |
| No | 5,984 (83.8%) |  | 273 (83.5%) |  | 49 (77.8%) |  |
| Yes | 874 (12.2%) |  | 38 (11.6%) |  | 11 (17.5%) |  |
| **Household Alcohol Consumption Rules** |  | <0.001 |  | 0.238† |  | 0.890† |
| Not Allowed | 5,628 (78.8%) |  | 247 (75.5%) |  | 46 (73.0%) |  |
| Allowed | 72 (1.0%) |  | 1 (0.3%) |  | 1 (1.6%) |  |
| No Rules | 1,435 (20.1%) |  | 79 (24.2%) |  | 16 (25.4%) |  |
| **Family Conflict Score** |  | 0.276 |  | 0.078 |  | 0.094 |
| Mean (SD) | 2.02 (1.93) |  | 1.75 (1.81) |  | 2.48 (2.40) |  |
| **Alcohol Availability** |  | <0.001 |  | 0.105 |  | 0.076 |
| Easy | 1,782 (25.0%) |  | 120 (36.7%) |  | 19 (30.2%) |  |
| Difficult | 5,000 (70.0%) |  | 189 (57.8%) |  | 41 (65.1%) |  |
| **Peer Alcohol Use** |  | <0.001 |  | 0.918 |  | 0.678† |
| No | 6,765 (94.7%) |  | 313 (95.7%) |  | 62 (98.4%) |  |
| Yes | 117 (1.6%) |  | 12 (3.7%) |  | 1 (1.6%) |  |
| **Positive Alcohol Expectancies Score** |  | <0.001 |  | 0.134 |  | 0.231 |
| Mean (SD) | 5.61 (2.61) |  | 6.06 (2.66) |  | 5.90 (2.59) |  |
| **Negative Alcohol Expectancies Score** |  | 0.098 |  | 0.238 |  | 0.636 |
| Mean (SD) | 12.1 (3.13) |  | 12.4 (2.64) |  | 12.4 (2.93) |  |
| **Neighborhood Safety Score** |  | 0.001 |  | 0.601 |  | 0.845 |
| Mean (SD) | 3.85 (0.99) |  | 3.98 (0.95) |  | 3.93 (0.98) |  |
| † Denotes use of Fisher's Exact Test. Otherwise, chi-squared tests were used for categorical variables, and t-tests were used for continuous variables.  Note: Percentages may not sum to 100% due to missing data. | | | | | | |
